# Electrical Stimulation for the Induction of Seizures is Safe, Well-tolerated and High-Yield in Children with Drug-Resistant Epilepsy

**DOI:** 10.1101/2025.06.17.25329709

**Authors:** Lucy Tarcha, Darcy A. Krueger, Hansel M. Greiner, Jesse Skoch, Francesco T. Mangano, Ravindra Arya, S. Katie Z. Ihnen

**Affiliations:** University of Cincinnati College of Medicine; Division of Neurology, Cincinnati Children’s Hospital Medical Center; Department of Pediatrics, University of Cincinnati College of Medicine; Division of Pediatric Neurosurgery, Cincinnati Children’s Hospital Medical Center

**Keywords:** drug-refractory epilepsy (DRE), children, electrical stimulation, epilepsy surgery, stereo electroencephalography (sEEG)

## Abstract

**Introduction:** One third of children with epilepsy develop drug-resistant epilepsy (DRE). Epilepsy surgery is the treatment for DRE, and selecting the surgical target typically requires recording spontaneous seizures during intracranial monitoring with stereoelectroencephalography (sEEG). In adults, induced seizures have been shown to help define surgical targets and are associated with superior surgical outcomes. No published studies focus on the safety, tolerability, and yield of electrical stimulation for the induction of seizures (ESIS) in children.

**Methods:** Stimulation at 1 Hz and 50 Hz was performed prospectively and comprehensively (all gray matter channels) during sEEG in children and young adults with DRE. Induced seizures were categorized into four groups: electroclinical habitual, clinical habitual (aura), subclinical and non-habitual. Primary safety outcome was use of rescue medication. Patient and caregiver tolerability were assessed by questionnaire. Yield was measured as induced seizure rate.

**Results:** Sixty-seven patients (n=30 female) were included, ages 1-21 years (mean ± SD, 11.1 ± 5.7). Three patients participated twice, yielding 70 patient admissions. n=10,135 stimulation trials were performed, requiring 175 ± 61 minutes per patient. Only one seizure required acute rescue medication. Patient and caregiver tolerability ratings were overall favorable and improved from pre- to post-session. At least one induced seizure was obtained in 62/70 (89%) patients, representing 283/10,135 trials (2.8%). Combined, habitual seizures and habitual auras accounted for most induced seizures (58%); non-habitual seizures were uncommon (12%). Of those with induced seizures, multiple seizure types occurred in 28/62 (45%). Seizures were induced at both 1 Hz and 50 Hz in 22/67 patients (33%).

**Significance:** Comprehensive seizure stimulation during sEEG is safe, well-tolerated, and high-yield in children. Both 1 Hz and 50 Hz stimulation induce seizures. These findings motivate a more systematic utilization of ESIS in pediatric EMUs.

## Introduction

Drug-refractory epilepsy (DRE) ^1^ afflicts one third of patients with epilepsy^2^, representing a significant^3,4^ and costly^5^ public health burden. The treatment for DRE is epilepsy surgery but its success varies from 30-70%^6–12^, mostly as a function of how accurately the surgical target is identified.

Many epileptologists use the seizure onset zone (SOZ) ^13^ as the surgical target. Localizing the SOZ is challenging in children due to multifocal abnormalities on non-invasive studies^12,14^ and seizure semiologies that may be subtle or poorly reported^15^ due to age or delays^16^. Invasive monitoring using stereoencephalography (sEEG) is thus increasingly used by pediatric epileptologists to identify the SOZ^17,18^. The gold standard for SOZ definition is the earliest EEG change before the clinical onset of the patient’s habitual (i.e., typical) seizure^13^; *several* seizures are required to confirm the consistency of the SOZ localization. Capturing multiple seizures may take days or weeks^19,20^, incurring cost^21^ and potential morbidity^22^. A technique that is safe, well-tolerated and high-yield for confirming SOZ localization in children with DRE – as an *adjunct* to the gold standard – is therefore of high value from both clinical and resource utilization perspectives.

Electrical stimulation for the induction of seizures (ESIS) was pioneered more than a century ago^23–25^. When Bancaud and Talairach developed sEEG^26–30^, they presented seizure stimulation as a fundamental part of the methodology^31^. Subsequent studies corroborated Bancaud and Talairach’s premise that induced seizures are integral to localizing the surgical target^32–34^. Contemporary French guidelines on sEEG^35^ continue to promote ESIS as part of sEEG monitoring, and 2025 American Clinical Neurophysiology Society (ACNS) guidelines also encourage the practice^36^. Despite this, many pediatric epilepsy monitoring units (EMUs), particularly in North America, perform ESIS in an *ad hoc* fashion at best^17,37^. Concerns about patient safety and tolerability have likely contributed to the limited implementation of ESIS in children.

The underutilization of ESIS in children reflects the lack of age-specific guidance in the literature. Older^32–34^ and newer^19,38–42^ research articles – as well as meta-analyses^43^ and reviews^44^ – have shown that induced seizures are safe, electroclinically similar to spontaneous seizures and associated with superior surgical outcomes. However, this literature focuses almost exclusively on adults, aside from a few reports describing small pediatric cohorts^45^ and mixed adult/child cohorts^46–48^ including a limited number of older children. *No published data focuses specifically on how to perform and interpret ESIS in children.* The primary aim of this prospective study was to investigate the safety, tolerability, and yield of ESIS in children and young adults with DRE.

## Methods

### Patient Sample

This prospective study was approved by the Cincinnati Children’s Hospital Medical Center (CCHMC) Institutional Review Board (IRB). Patients undergoing sEEG monitoring were recruited consecutively from the CCHMC EMU from February 2022 – January 2025. Minimum age was 1 year. Clinically unstable patients and those whose seizures always require rescue were excluded. Patients and/or caregivers provided written informed consent, and children over age 10 years provided assent. A detailed epilepsy history was obtained for each patient.

### Stereo EEG Implantation and Signal Recording

Pre-surgical electrode planning was determined by multi-disciplinary consensus following non-invasive evaluation. No additional electrodes were inserted for this study. Contacts distal from the hypothesized SOZ and minimally affected by artifact were chosen for reference and ground. sEEG electrodes had cylindrical recording contacts with 0.8 mm diameter, 2 mm contact length, and 4.3 mm center-to-center distance (PMT Corporation, Chanhassen, MN). EEG was sampled at 2048 Hz using Natus (n=59; Natus Medical Incorporated) and Cadwell Arc 2.2 (n=11; Cadwell Industries, Incorporated) software.

### Tolerability

Tolerability was initially assessed via a survey administered immediately before and again after stimulation. One question asked, “Do you have any concerns about participating in this study?” and another asked, “What is your level of comfort with participating in this study?”. Starting with patient 26, 10-point Likert scales were attached to each question to increase objectivity and allow for responses to be quantified. As able, both patient and caregiver(s) completed both the pre- and post-questionnaires.

### Stimulation Approach and Planning

Cortical stimulation was performed at least 24 hours after sEEG implantation to avoid post-operative confounds including anesthesia ^44,49^. Sessions were held during the day with the patient awake, whenever possible, to limit state-dependent variations in cortical excitability ^50^. Bipolar stimulation with cathode-leading ^51^ biphasic square-wave pulses^52,53^ targeted pairs of adjacent electrode contacts (“channels”) in gray matter ^54^. Channels outside brain or with abnormal impedances or prevalent artifact were excluded.

Each patient’s stimulation plan was custom-made based on the predicted relevance of each channel for the SOZ, informed by non-invasive data and interictal sEEG activity. Channels predicted most likely to be involved in the SOZ were generally stimulated last.

### Stimulation Paradigm

Electrical stimulation was performed using a systematic approach based on published parameters ^52,55,56^ consistent with ACNS technical standards^36^ and modified based on strategies reported in adults ^43,57^. Stimulation at all gray matter channels using low frequency (1 Hz) was performed first, followed by stimulation at all gray matter channels using high frequency (50 Hz). EEG activity was monitored at the bedside by a single fellowship-trained epileptologist (SKZI) to ensure return to baseline^58^ between stimulations. The minimum stimulation-end to stimulation-start interval was 30 seconds. Refractory period effects ^57^ were further avoided by not stimulating consecutively at adjacent contacts. If motor function was induced at 1 Hz at a particular channel, stimulation at 50 Hz was not performed there.

### Stimulation Parameters

For both frequencies of stimulation, current ranged from 1-10 mA and pulse width from 250-300 microseconds. Maximum charge density was 57µC/cm²^51,59^. Train durations were 30 and 5 seconds for 1 and 50 Hz stimulations, respectively.

A “global” current intensity threshold was estimated for each patient at the beginning of low frequency and again at the start of high frequency stimulation. Stimulation began at 1 mA and amperage was increased stepwise (by 0.5-1 mA) on successive trials (at different channels) until we obtained a) after-discharges (evoked electrographic changes without evolution); b) a clinical response (typically sensorimotor, including pain/discomfort); c) maximum charge density or d) an induced seizure. Channels initially stimulated at intensities below threshold were stimulated again later at threshold, so that each channel was given an equal “chance” to have an induced response. Due to regional variability in cortical excitability^60,61^, the “global” threshold was additionally titrated throughout the session^62^. I.e., if stimulation at a channel did not result in one of the above-mentioned endpoints, that channel was stimulated *later* at an amperage 0.5– 1 mA higher. In regions with high cortical excitability (e..g, mesial temporal and perirolandic), stimulation started *below* the “global” threshold and likewise was increased as required to reach one of the above-stated endpoints.

### Possible Outcomes of Each Stimulation

Stimulation ceased at a channel if any above-listed endpoint was reached. If a seizure occurred during ESIS, whether induced or spontaneous, stimulation was paused until the patient was back to baseline clinically and per the EEG. Stimulation then proceeded at the next (non-adjacent) channel. Patients could have induced seizures from more than one channel.

### Seizure Rescue During ESIS

If a patient had a prolonged induced seizure of generalized convulsive semiology (>5 minutes), the seizure was treated according to the patient’s clinical rescue plan and ESIS ended.

### Data Extraction

All completed trials were analyzed. For each induced seizure, electroclinical features were examined by a board-certified epileptologist (SKZI), including semiology, morphology of EEG onset ^63–65^, EEG spread within 3 seconds ^57,66,67^, recruited ictal patterns, and seizure duration. Induced seizures were compared to spontaneous seizures (SpSz) using these criteria and categorized as follows: electroclinical habitual (semiology and EEG similar to SpSz), clinical habitual (early SpSz semiology with or without EEG change), subclinical (no clinical change with EEG evolution), and non-habitual (atypical semiology with EEG evolution). See **Figure 1** for examples of each type of seizure. All four of these categories are distinguished from after-discharges because they evolve, as per ACNS criteria^68^.

**Figure 1.**
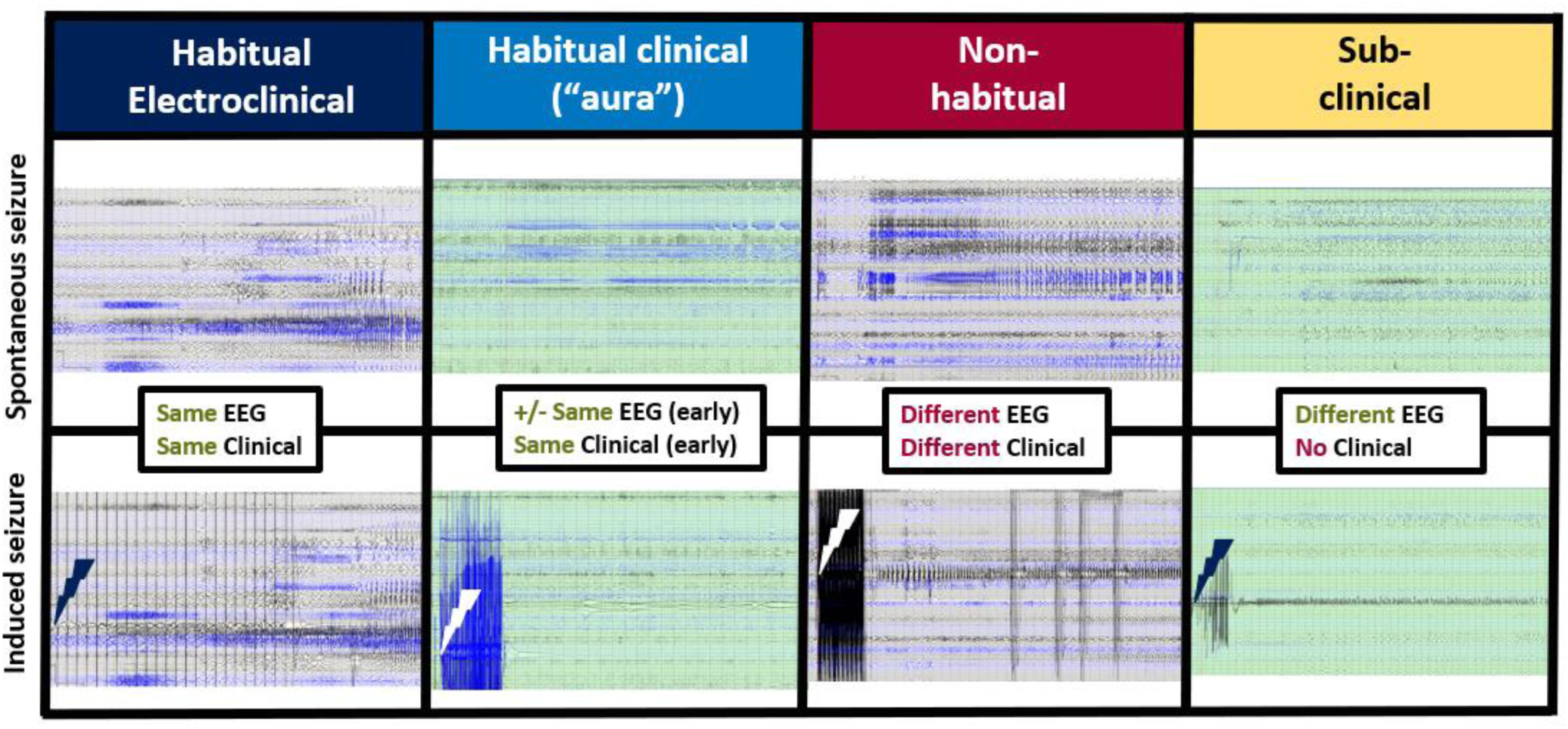
Examples of the four types of seizures that were induced. Each column shows an example of one patient’s habitual electroclinical seizure (“spontaneous seizure”) in the top panel, along with an induced seizure from that same patient in the bottom panel. The lightning strike points to the electrode channel that was stimulated in that trial. Each example was chosen from a different patient; the electroclinical habitual seizure was induced by low frequency stimulation and the others were induced by high frequency stimulation. Stimulation artifact for both frequencies can be seen on the tracings, as can artifact from four pulses of ictal disrupt applied during the non-habitual seizure. **Habitual electroclinical** induced seizures are similar to spontaneous seizures both electrographically (EEG) and clinically. **Habitual clinical** induced seizures (“auras”) recapitulate early clinical features of the spontaneous seizures and a variable degree of early development on EEG. **Nonhabitual** induced seizures have an electrographic pattern and clinical features that are both atypical (different from the patient’s spontaneous seizures). **Subclinical** induced seizures have an evolving electrographic change (per ACNS criteria^68^) without any discernible clinical change. Note that induced subclinical seizures may or may not resemble an electrographic pattern seen spontaneously, since some patients have spontaneous subclinical seizures, but the EEG pattern is (by definition) not the EEG pattern seen with the patient’s spontaneous habitual electroclinical seizures. Subclinical seizures are *not* synonymous with after-discharges, which do not evolve.

### Statistical Analysis

Statistical analyses were conducted using Python. Continuous variables were compared between groups using *t*-tests and ANOVAs. Group differences in the observed proportions of dichotomous categorical variables with small numbers of observations were evaluated using Fisher Exact test; for categorical variables with more than two categories and for those with large numbers of observations, Chi-squared test was used. A general linear mixed model was used to evaluate interactions between continuous and categorical variables at the level of the trial. Pearson correlation coefficient was used to measure the strength and direction of the relationship between two continuous variables. For all tests, a two-sided *p*<0.05 was considered statistically significant.

## Results

### Demographic Data

During the study period, n=95 sEEG patients met inclusion criteria; an additional n=6 sEEG patients could not be offered participation due to schedule conflicts with study personnel. Seventy-two of 95 patients enrolled, representing a 76% enrollment rate. These analyses focus on patients ages 21 years and younger, which excludes n=2; thus the analyzed cohort includes n=70 complete stimulations in n=67 participants (n=30 female), three of whom (n=1 female) participated twice. The time between sEEG admissions for the patients who participated twice was 28-266 days.

**Table 1** summarizes demographic and stimulation session characteristics. The mean age during ESIS was 11.1±5.7 years. Patients in whom seizures could be induced tended to be older than those in whom seizures were not induced (mean 11.1 vs. 7.7 years); however this difference was not significant (**Table 1**). Fifty-six patients (80%) had a lesion on MRI, of whom 21 (30% of total cohort) had Tuberous Sclerosis Complex (TSC). Other lesional etiologies included focal cortical dysplasia, birth-related injuries including perinatal strokes and brain tumors. The sEEG implants were left hemisphere in 27 patients (38.5%), right in 20 (28.5%), and bilateral in 23 (33%). All 20 patients (100%) with unilateral right hemisphere implants had at least one induced seizure, whereas only 21/27 (78%) of those with unilateral left hemisphere implants had at least one induced seizure, a significant difference (statistics in **Table 1**).

**Table 1.** Patient and session characteristics, stratified by whether the patient had at least one seizure induced (of any kind) for continuous variables, mean (SD); for categorical variables, number (proportion)

| Patient and Session Characteristics, Continuous |  |  |  |  |  |  |  |
| --- | --- | --- | --- | --- | --- | --- | --- |
|  | Overall Cohort (n=70) | Seizure Induced (n=62) | No Seizure Induced (n=8) | p value | Hedge's g | t statistic |  |
| Age at Stimulation, years | 11.1 (SD 5.7) | 11.4 (SD 5.8) | 7.7 (SD 5.2) | 0.094 | -0.6299 | -1.6956 |  |
| Age of Epilepsy Onset, years | 4.4 (SD 4.4) | 4.6 (SD 4.4) | 3.1 (SD 4.7) | 0.38 | -0.3254 | -0.8758 |  |
| Duration of Epilepsy, years | 6.6 (4.2) | 6.8 (SD 4.3) | 4.6 (SD 3.2) | 0.17 | -0.5348 | -1.4395 |  |
| Duration of sEEG, nights | 6.2 (SD 5.0) | 6.2 (SD 5.2) | 5.5 (SD 3.5) | 0.69 | -0.1480 | -0.3985 |  |
| Number of electrodes | 14.3 (SD 3.9) | 14.3 (SD 4.0) | 14.0 (SD 4.0) | 0.83 | -0.0802 | -0.2159 |  |
| Number of electrode contacts | 158 (SD 48) | 160 (SD 49) | 145 (SD 46) | 0.43 | -0.2980 | -0.8022 |  |
| Days between sEEG implant and stim. | 2.9 (SD 2.9) | 3.0 (SD 3.0) | 2.8 (SD 3.1) | 0.86 | -0.0678 | -0.1825 |  |
| Patient and Session Characteristics, Categorical |  |  |  |  |  |  |  |
|  | Overall Cohort (n=70) | Seizure Induced (n=62) | No Seizure Induced (n=8) | p value (two-sided) | Test statistic | Diff. (Δ) in proportions | Wald 95% CI for Δ, with Yates correction |
| Sedated on dexmedetomidine in ICU |  |  |  |  |  |  |  |
| Yes | 4 | 3 (0.75) | 1 (0.25) | 0.39 | 0.36 (FET) | 0.14 | (-0.42, 0.70) |
| No | 66 | 59 (0.89) | 7 (0.11) |  |  |  |  |
| Spontaneous seizure <24 h prior to stimulation |  |  |  |  |  |  |  |
| Yes | 47 | 42 (0.89) | 5 (0.11) | 1.0 | 1.26 (FET) | 0.02 | (-0.17, 0.22) |
| No | 23 | 20 (0.87) | 3 (0.13) |  |  |  |  |
| Spontaneous seizure <24 h after stimulation |  |  |  |  |  |  |  |
| Yes | 44 | 39 (0.87) | 5 (0.13) | 0.80 | 1.02 (FET) | 0.002 | (-0.18, 0.18) |
| No | 26 | 23 (0.88) | 3 (0.12) |  |  |  |  |
| All spontaneous seizures occurred AFTER stimulation |  |  |  |  |  |  |  |
| Yes | 8 | 7 (0.875) | 1 (0.125) | 1.0 | 0.89 (FET) | 0.01 | (-0.32, 0.30) |
| No | 62 | 55 (0.89) | 7 (0.11) |  |  |  |  |
| Laterality of sEEG implant* |  |  |  |  |  |  |  |
| Right | 20 | 20 (1.0) | (0.0) | 0.03 | inf.* (FET) | 0.22 | (0.02, 0.42) |
| Left | 27 | 21 (0.78) | 6 (0.22) |  |  |  |  |
| Bilateral | 23 | 21 (0.91) | 2 (0.09) |  |  |  |  |
| **Medication Status\*\*** |  |  |  |  |  |  |  |
| On (full dose) | 17 | 15 (0.88) | 2 (0.12) | 0.94 | 0.007 ($\chi^2$) | 0.006 | (-0.183, 0.196) |
| On (reduced dose) | 9 | 8 (0.89) | 1 (0.11) |  |  |  |  |
| Off | 41 | 36 (0.88) | 5 (0.12) |  |  |  |  |
| Mixed | 3 | 3 (1.0) | 0 (0.0) |  |  |  |  |
| **Sex** |  |  |  |  |  |  |  |
| Female | 31 | 28 (0.90) | 3 (0.10) | 1.0 | 1.37 (FET) | 0.03 | (-0.15, 0.21) |
| Male | 39 | 34 (0.87) | 5 (0.13) |  |  |  |  |
| **Handedness\*\*\*** |  |  |  |  |  |  |  |
| Right-handed | 46 | 42 (0.91) | 4 (0.09) | 1.0 | 1.31 (FET) | 0.02 | (-0.18, 0.23) |
| Left-handed | 18 | 16 (0.89) | 2 (0.11) |  |  |  |  |
| *Ambidextrous/Unknown* | 6 | 4 (0.67) | 2 (0.33) |  |  |  |  |
| **MRI** |  |  |  |  |  |  |  |
| Lesional | 56 | 49 (0.88) | 7 (0.12) | 1.0 | 0.54 (FET) | 0.05 | (-0.26, 0.15) |
| Non-lesional | 14 | 13 (0.93) | 1 (0.07) |  |  |  |  |
| **Prior Epilepsy Surgery** |  |  |  |  |  |  |  |
| Yes | 14 | 12 (0.86) | 2 (0.14) | 0.66 | 0.72 (FET) | 0.03 | (-0.28, 0.20) |
| No | 56 | 50 (0.89) | 6 (0.11) |  |  |  |  |

Counts represent 70 patient sEEG admissions, including three patients who had two distinct sEEG admissions.
FET = Fisher exact test; $\chi^2$ = Chi-squared test.
\*For comparing sEEG laterality groups, the right and left sided implants were compared using Fisher Exact Test, and the bilateral group was ignored. Infinity resulted as the test statistic because there were zero observations of “no seizure induced” in the group with right-sided sEEG implants. This is an actual test result, not an error. When a Chi-squared test was computed on the 3x2 contingency table that included those with bilateral implants, the resulting $\chi^2$ test statistic was 5.86 and the two-sided $p$ value was 0.053, approaching significance. This suggests that even when including the bilateral implants, there is an effect of hemisphere of implantation on the likelihood that a patient will have an induced seizure.
\*\* For ASM status, On (full dose) and On (reduced dose) were combined and compared against Off. The 3 patients with mixed medication status (on meds for some trials, off meds for others) were ignored.
\*\*\* Similarly, for handedness, right and left handed patients were compared using Fisher Exact Test, the small number of patients (n=6) in the ambidextrous/unknown group were ignored.

### Characteristics of Stimulation Sessions

The time between sEEG implant and the start of stimulation was 2.9±2.9 days. The mean total run time per patient was 175±61 minutes. Stimulations were conducted over one (n=48), two (n=20) or three days (n=2). On average, patients underwent 79±25 low frequency and 68±33 high frequency trials. The number of 50 Hz trials was overall lower than the number of 1 Hz trials due to our approach of skipping 50 Hz stimulation for channels at which 1 Hz stimulation induced a sensorimotor change (see methods). In total, 10,135 ESIS trials were performed, with 5,929 (59%) targeting left-sided channels and 4,206 (41%) targeting right-sided channels. High-frequency stimulations accounted for 4,796 (47%) trials, while low-frequency stimulations comprised 5,339 (53%).

### Safety Outcomes

Of 283 induced seizures in 70 patients, only one seizure in one patient required acute rescue medication (primary safety endpoint). This patient was rescued at two minutes in accordance with her home plan. Six additional patients experienced non-habitual motor seizures that generalized but did not require rescue (secondary safety endpoint). One of these 6 patients requested to end his stimulation session thereafter. All seven of these patients were teenagers and young adults, with a mean age of 19.2 (range 15.7-21.1) years. Four were MRI non-lesional. High-frequency stimulation induced six of the seven seizures.

Paresthesias and/or clonic jerking were induced *during* stimulation in nearly all patients who had electrodes in sensorimotor cortex, as expected. Four patients additionally reported having (or were perceived by caregivers to have had) unexpected, unpleasant sensations induced by ESIS. Examples included a brief stinging sensation near the scalp (likely meningeal stimulation) and a loud noise (likely primary auditory cortex stimulation). These phenomena did not outlast the stimulus duration.

### Tolerability Outcomes

Prior to ESIS, patients and caregivers expressed various concerns (see **Appendix A** and **Appendix B**). Patients were primarily worried about what the test might feel like. Caregivers were concerned about possible pain and often mentioned wanting to avoid induced seizures of atypical semiology. They expressed fears about prolonged induced seizures and the potential long-term effects of “extra” seizures. Regarding comfort before the session, patients generally described themselves as “a little nervous.” Caregivers expressed a desire to ensure that no unknown factors were introduced that could impact the overall sEEG interpretation. Despite their concerns, caregivers expressed that they generally trusted the medical team.

After the test, patient concerns decreased, with many reporting “no concerns” and finding the process “very easy.” Caregiver concerns varied; some were worried about seizures occurring in different locations, while others noted that seizures subsided faster than expected and the stimulation duration was shorter than anticipated. Patients generally rated their post-test comfort positively. One patient found the progressive ramp-up to simulate seizures “very informative.” Caregiver comfort levels varied, with one remarking that the experience was “worth the risk.” Another caregiver mentioned feeling comfortable because rescue medications were available.

Pre- and post-stimulation ratings for concerns and comfort level were quantified for the subset of respondents: caregivers (n=44) and patients (n=21). For both groups for both items, median scores improved (became more positive) from pre- to post-stimulation, as shown in **Figure/Table 2**. For caregivers, median ratings went from 2 to 1 for concerns and from 4 to 2 for comfort; for patients, from 2 to 1 for concerns and from 4 to 3 for comfort. For the caregivers, the differences between the pre- and post-scores were significant (**Figure/Table 2**).

**Figure/Table 2.**
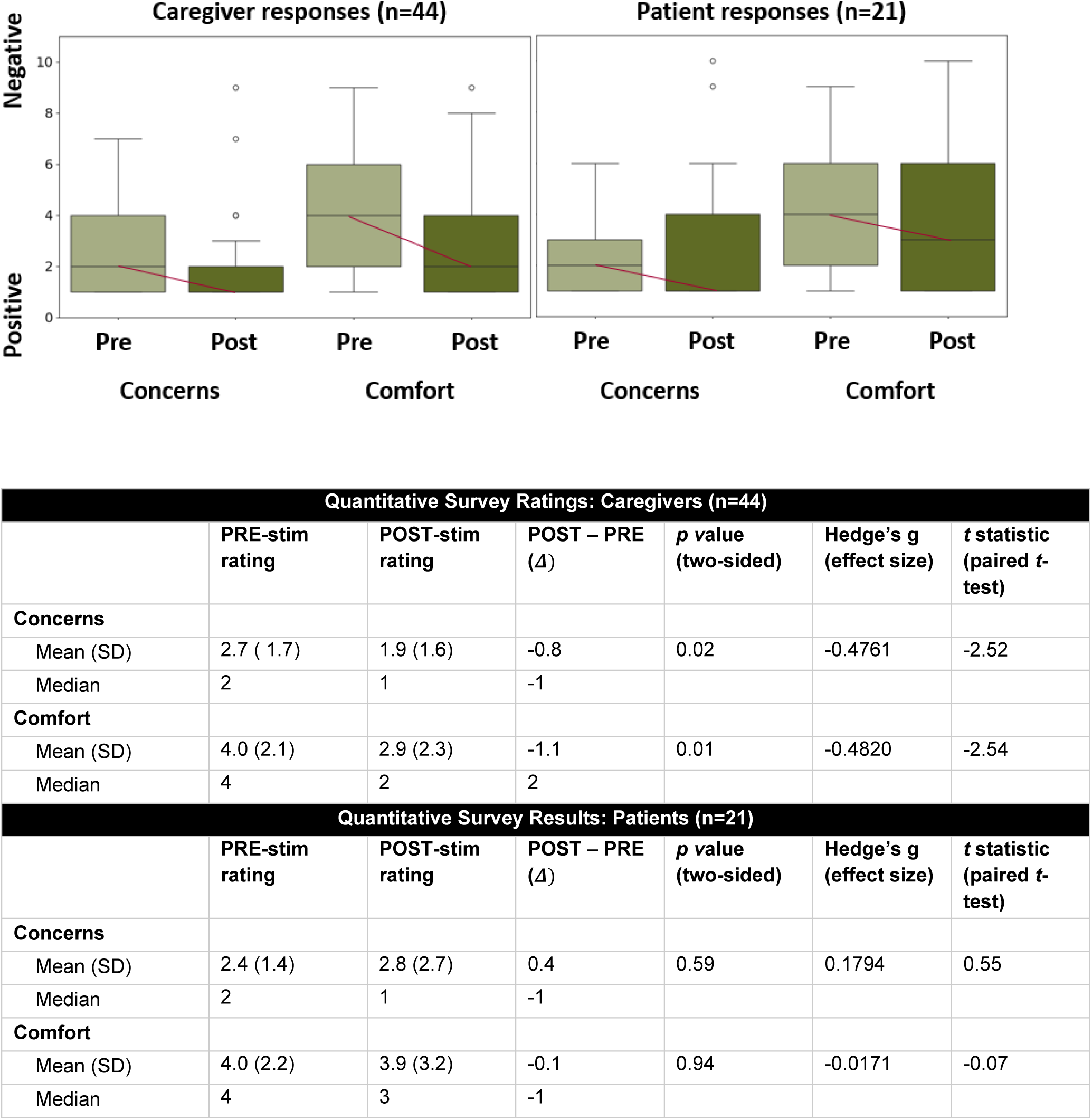
Ratings of comfort and concern for caregivers and patients. n=44 caregivers and n=21 patients responded. The response range included the integers from 1 to 10, with lower numbers reflecting more positive ratings (i.e., fewer concerns or greater comfort). Caregiver responses are shown in the left panel **(Panel A)** and patient responses are shown in the right panel **(Panel B)**. Pre indicates ratings before the stimulation began, while Post indicates ratings after the conclusion of the stimulation.

### Yield of Seizure Induction

Two levels of observation are relevant in this study: the individual patient and the individual trial (with patient as a nested variable).

### Overall Yield

#### Level of the individual patient

Of n=70 patients, 62 (89%) experienced *at least one* induced seizure, and 46 (66%) specifically had *at least one* induced habitual seizure (electroclinical or clinical “aura”) (**Figures 3A, 3B)**. Among 62 patients who could be induced, the mean number of induced seizures was 4.6 (range 1-21; median 3). **Figure 3C** shows what proportion of patients experienced at least one of each induced seizure type. Habitual electroclinical seizures were induced in the largest proportion of patients, 30/70 (0.42), while subclinical seizures were induced in 24/70 (0.34), nonhabitual seizures in 23/70 (0.33) and habitual auras in 18/70 (0.26). As shown in **Figure 3D**, the plurality of patients (28/70, or 0.40) had more than one type of induced seizure (for example, a habitual electroclinical induced seizure and a subclinical induced seizure). Fifteen patients of 70 (0.21) had exclusively habitual electroclinical induced seizures; seven (0.10) had exclusively non-habitual seizure and six (0.09) each had exclusively habitual clinical (“aura”) or exclusively subclinical induced seizures. Notably, in 22 of the 67 patients who had at least one induced seizure (0.33), seizures were induced at both high and low frequency (often at different channels).

**Figure 3.**
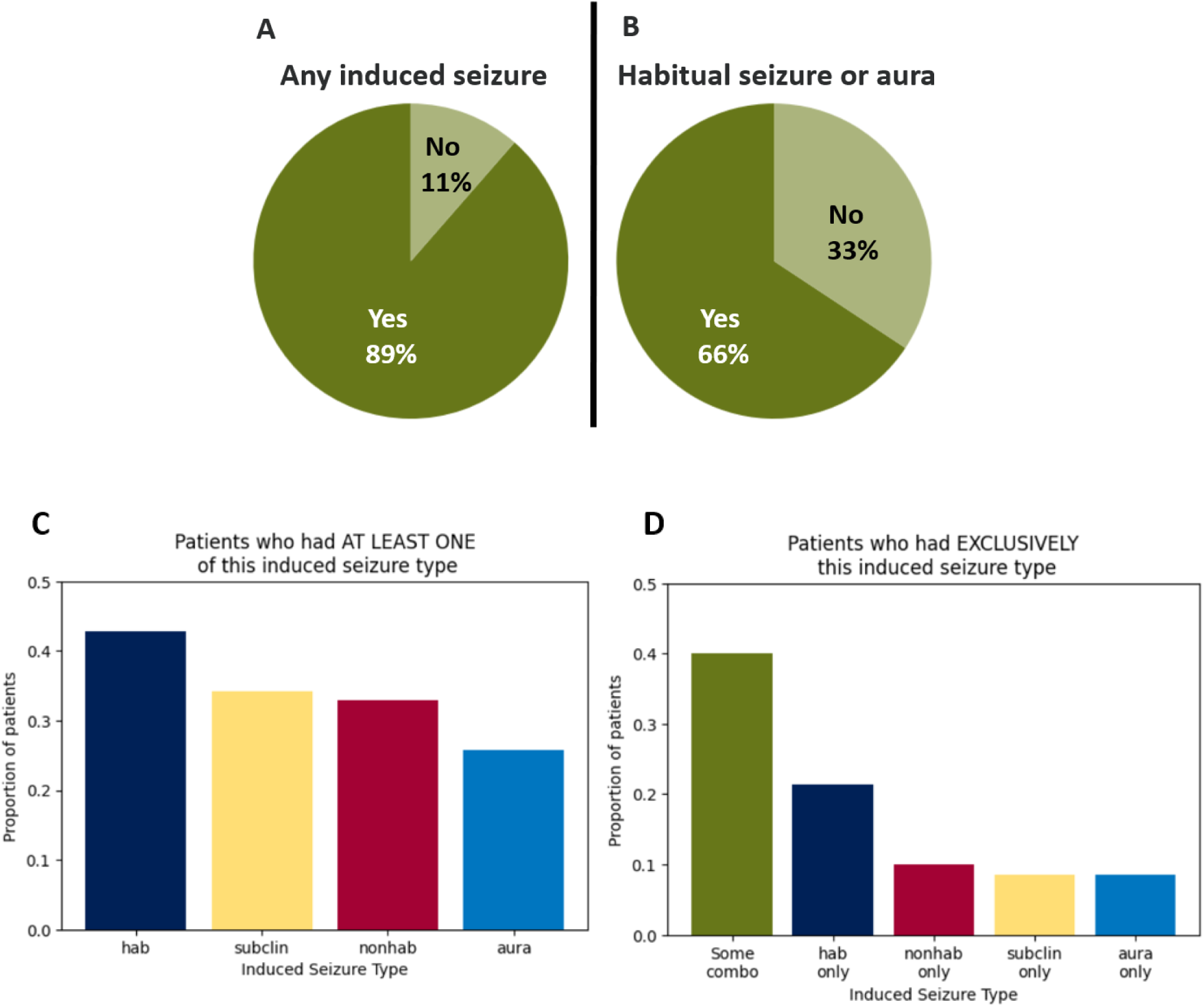
Stimulation yield at the level of the individual patient. **Panel A** shows that 89% of all patients had at least one induced seizure, and **Panel B** shows that 66% of patients had at least one induced habitual eletroclinical *or* habitual clinical (“aura”) seizure. In **Panel C**, the proportion of patients who had at least one of each of the four induced seizure types is plotted, whereas in **Panel D**, the proportion of patients who had exclusively type of induced seizure is shown. The majority of patients had some combination of induced seizure types. hab = habitual electroclinical seizure; subclin = subclinical induced seizure; nonhab = nonhabitual induced seizure and aura = habitual clinical (“aura”) induced seizure.

In contrast to prior observations in adults^40^, whether or not the patient had a spontaneous seizure in the 24 hours prior to ESIS did not affect whether or not a seizure could be induced. **Table 1** shows that several other features did *not* significantly differ between patients who had at least one induced seizure (n=62) and those who did not (n=8), including presence of MRI lesion and anti-seizure medication (ASM) status. **Supplemental table 1** is analogous to Table 1 but divides the sample into those who had at least one *habitual* induced seizure (n=46) vs. those who did not (n=24). None of the examined factors differed between those two sub-groups.

#### Level of the individual trial

The total number of induced seizures was 283, representing 2.8% of 10,135 trials. The large denominator reflects our comprehensive approach of stimulating every gray matter channel. The most common induced seizures were habitual electroclinical (n=107; 38% of 283), followed by subclinical (n=85; 30%), habitual clinical (auras) (n=56; 20%), and non-habitual (n=35; 12%). The proportion of 50 Hz stimulations that resulted in an induced seizure was significantly greater than that of 1 Hz stimulations (0.037 vs. 0.020; **Table/Figure 4**). The same was true if only *habitual* induced seizures were considered (0.019 vs. 0.013; **Table/Figure 4**).

**Table/Figure 4.**
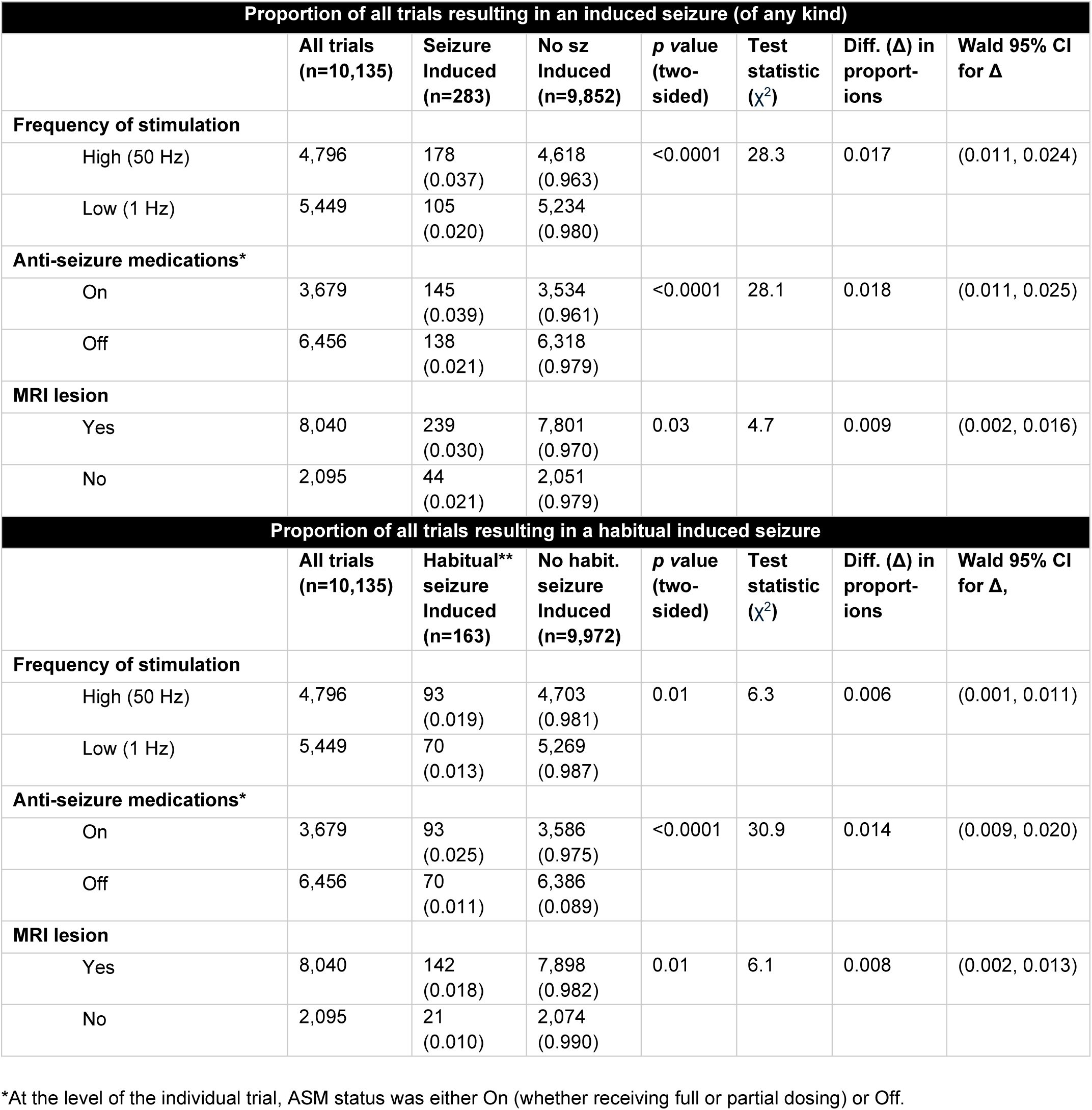
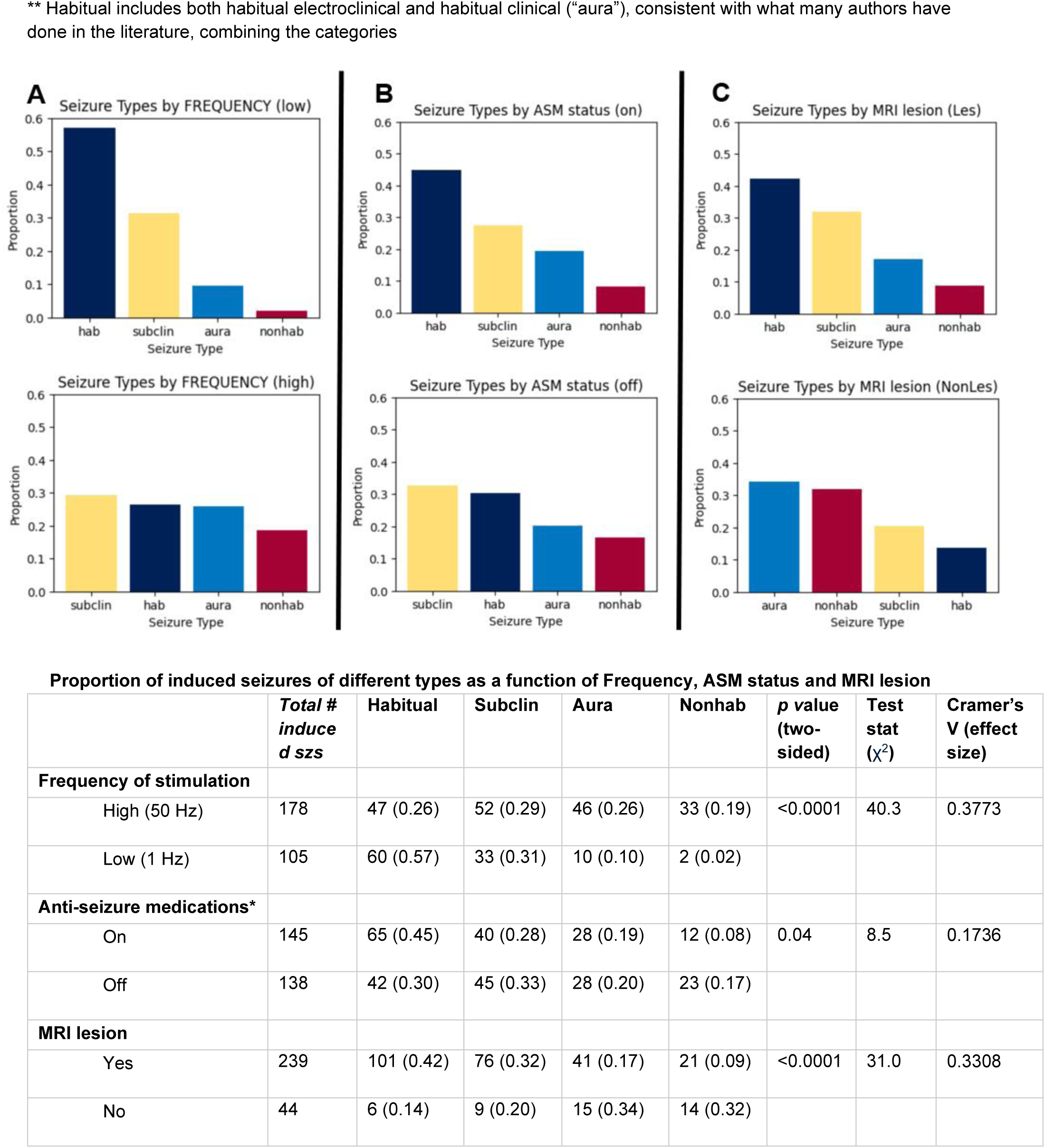
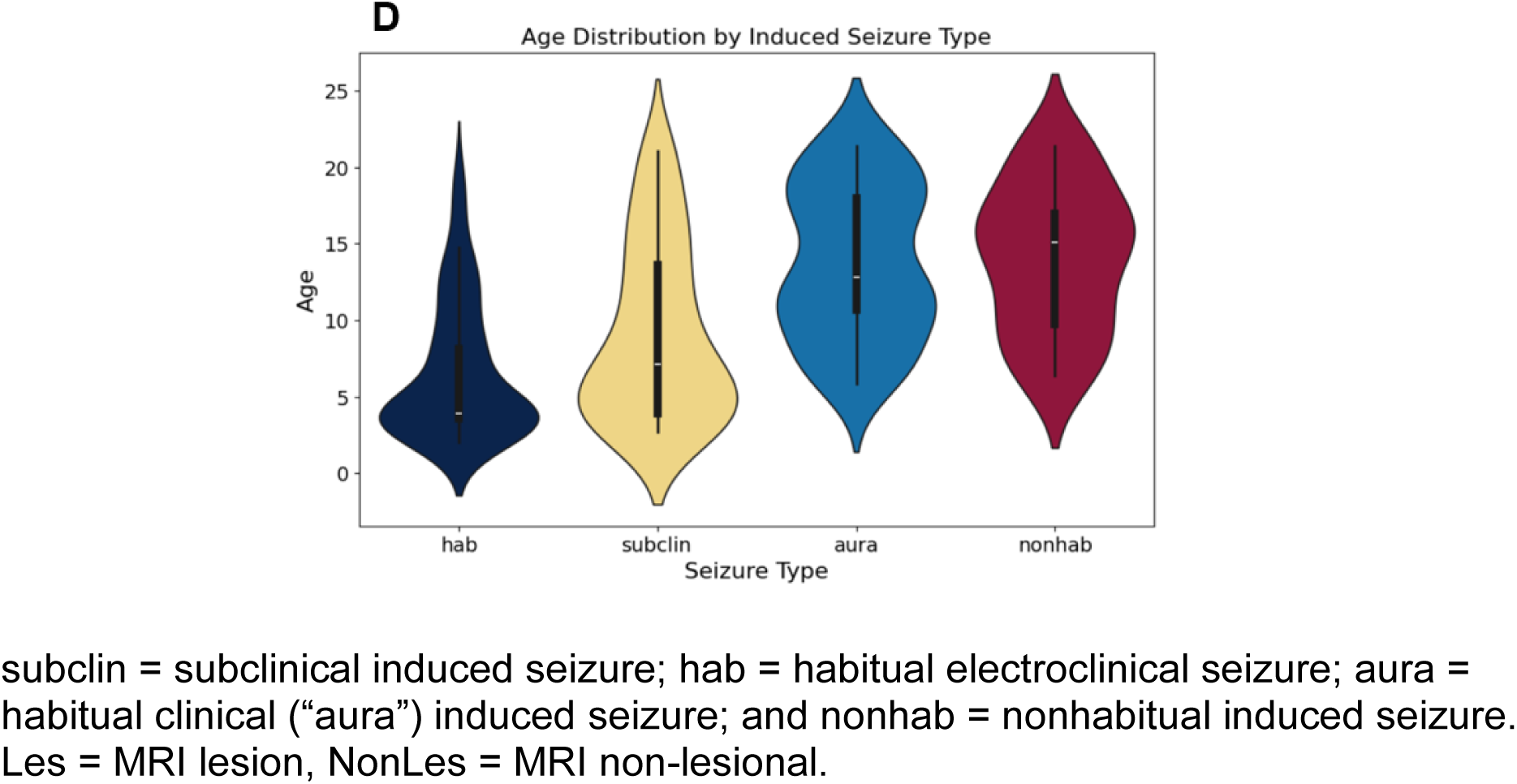
Stimulation yield at the level of the individual trial. The first **Table** shows the proportions of trials resulting in an induced seizure of any type (top) or habitual (bottom), as a function of frequency, anti-seizure medication (ASM) status and MRI lesion. The **Figure** and second Table show, for all n=283 induced seizures, the relative proportions of each type, again as a function of frequency **(A)**, ASM status, **(B)** and MRI lesion **(C)**. Y-axes are the same for A, B and C. **Panel D** shows the age distribution (across trials) of each induced seizure type.

As mentioned above and shown in **Table 1**, whether or not a patient was on ASMs at the time of ESIS did not affect whether that patient had *at least one* induced seizure. However, at the level of the individual trial, a significantly greater number of seizures were induced in patients *on* ASMs as compared to *off* (0.039 vs. 0.021; **Table/Figure 4**). The same was true if only *habitual* induced seizures were considered. In other words, patients on ASMs accounted for a greater number of induced seizures than those off ASMs.

#### Yield of Each Induced Seizure Type

The proportion of each type of induced seizure varied significantly by stimulation frequency. Specifically, the largest proportion of seizures obtained with low frequency stimulation were habitual electroclinical (0.57), whereas high frequency stimulation yielded a larger proportion of other seizure types (**Figure 4A**). Proportions of seizure types likewise varied significantly by ASM status, although the effect size was smaller. The largest proportion of seizures obtained in patients on ASM were habitual electroclinical (0.45), whereas subclinical and habitual electroclinical seizures were obtained in roughly equal proportions in patients off ASM (0.33 and 0.30; **Figure 4B**). The presence of an MRI lesion (not necessarily at the stimulated site) also significantly affected the proportions of induced seizure types. The largest proportion of seizures obtained in patients with MRI lesions were habitual electroclinical (0.42), whereas habitual auras and non-habitual seizures were observed in greater proportion in patients without MRI lesions (0.34 and 0.32; **Figure 54**). Regarding age, habitual electroclinical and subclinical seizures were relatively more prevalent in younger patients, as compared to non-habitual seizures and auras, which skewed older (*p*<0.0001; F-stat 34.95 by ANOVA; **Figure 4D**).

#### Age Effects

Additional age effects were noted. As shown in **Figure 5A**, age interacted significantly with frequency (*Z=*-5.44, *p*<0.000), such that younger children accounted for a greater proportion of the induced seizures at low frequency as compared to older children. The interaction was especially pronounced for those under age 5 years. Additionally, there was a significant negative linear relationship between charge density and age, whether this was examined overall (r(281)=-0.546, *p*=1.99e-23, R^2^=0.30; slope=-1.68) or separately for both high frequency (r(173)=-0.377, *p*=2.13e-07, R^2^=0.14; slope=-0.69) and low frequency (r(103)=-0.543, *p*=2.10e-09, R^2^=0.30; slope=-1.32). **Figure 5B** shows that younger patients required a higher charge density to induce seizures than older patients, particularly for low frequency. Also evident in **Figure 5B** is the observation that lower charge densities were required to induce seizures at high frequency as compared to low frequency (*t-*stat −21.6, *p*=8.85e-62).

**Figure 5.**
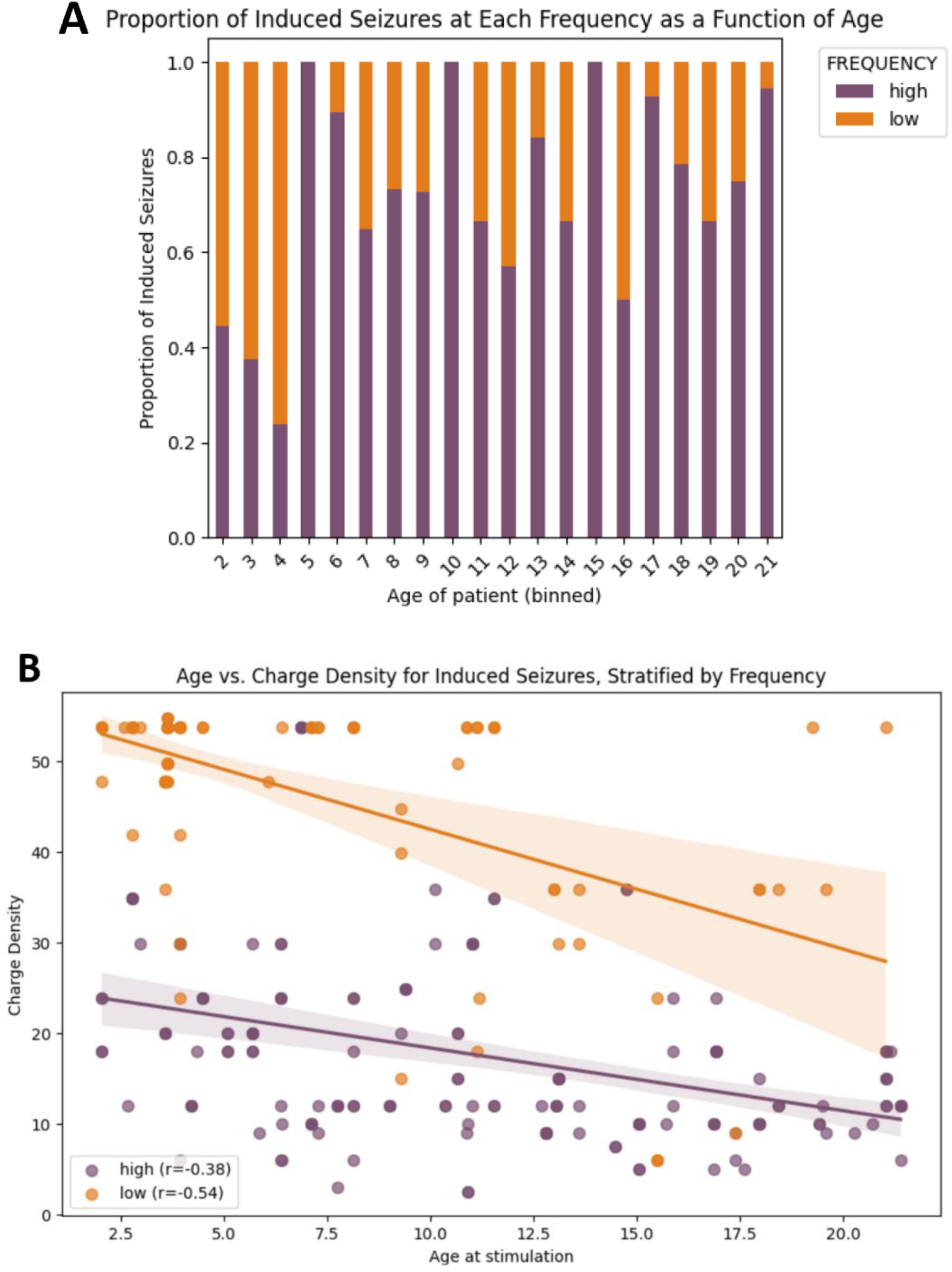
Age-related effects. **Panel A** shows the proportion of induced seizures obtained during high vs. low frequency across all trials at a given age (binned along the x-axis in whole years). Seizures at low frequency (orange), were disproportionately obtained in the youngest children (ages 2-4 years). **Panel B** shows age again along the x-axis, this time as a continuous variable, with each scatter plot representing a single induced seizure. Charge density is plotted along the y-axis. High and low frequency stimulations are color-coded separately. Linear regression lines are plotted for each frequency. For both frequencies, higher charge densities were required to induce seizures at younger ages.

## Discussion

Electrically induced seizures have been shown to be safe^40^ and informative for surgical planning in adults with DRE^41, 43^. In this first large prospective *pediatric* investigation of seizure stimulation, we show that ESIS using 1 and 50 Hz and performed comprehensively is likewise safe, well-tolerated and high-yield in a large single-center sample of n=70 children and young adults.

### Safety

The use of acute rescue medication was our primary safety endpoint. This choice was conservative, since it is not unusual for patients undergoing sEEG monitoring off ASMs to require rescue medication. Even so, our study showed that ESIS is safe in children and young adults: only one seizure in one patient required acute rescue, representing an adverse event rate at the patient-level of 1.4%. Six additional patients (9%) met our secondary safety endpoint (non-habitual motor seizures that generalized but did not require rescue), although only one asked to stop the stimulation thereafter. Notably, all seven of these patients were ≥15 years old, and all but one of these seizures was induced at high frequency.

Seventeen patients (about a quarter of our cohort) were 5 years old and younger, comprising an age group whose caregivers and physicians tend to be especially trepidatious about risk exposure. It was therefore especially heartening to confirm that none of the adverse events occurred in this sub-group of very young children.

### Safety: Suggestion for clinical practice

Patients and families can be counseled that for children and young adults, ESIS adds negligible additional risk beyond the risks inherent to sEEG monitoring. Furthermore, clinicians may choose to exercise extra caution when performing ESIS on teenagers and young adults (for example, by titrating current in smaller steps), particularly for stimulations at high frequency.

### Tolerability

Undergoing sEEG can be a stressful experience for patients and families, and clinician awareness of that fact is likely one reason that ESIS is not universally integrated into pediatric EMUs. However, these data affirm that ESIS in children and young adults is quite well-tolerated and even favorably viewed.

First, our study enrollment rate was 76%, which suggests that a thoughtful informed consent process goes a long way to pre-empt concerns. Additionally, patients and caregivers specifically had many positive comments about ESIS, with one dominant theme being the desire to help others. Importantly, ratings of comfort and concern showed improvements from pre- to post-ESIS, despite the long sessions (on average 3 hours). Of note, four patients completed the study while on dexmedetomidine for sedation due to behavioral challenges attributable to their underlying medical diagnoses, and even these children’s caregivers had satisfactory impressions of the process.

### Tolerability: Suggestions for clinical practice

These results demonstrate that children, teenagers and their caregivers tolerate seizure stimulation well, supporting systematic integration of ESIS into the workflow of the pediatric EMU.

### Yield

The proportion of patients with at least one induced habitual seizure (electroclinical or clinical) in this study was 66%. This is comparable to some prior reports^40,46,48,69^, but higher than was reported in one recent retrospective study focused on a small cohort of n=14 children^45^, of whom only n=4 (28%) had induced habitual seizures. Crucially, however, the participants in the Manokaran et al. study were selected for stimulation either because they were not having sufficient spontaneous seizures or because their SOZs were felt to be diffuse. Additionally, only n=3 of the patients underwent 50 Hz stimulation in addition to 1 Hz stimulation, whereas in our larger cohort, all patients underwent stimulation at both frequencies. (Of note, the Manokaran approach to using ESIS *selectively* in the pediatric EMU mimics common practice amongst pediatric epileptologists). Thus we attribute the notably larger yield in our study to our more systematic and comprehensive approach.

50 Hz stimulation was more effective than 1 Hz stimulation at inducing seizures overall, and this was true separately for non-habitual seizures and habitual seizures (combining across electroclinical habitual and clinical habitual). This is broadly consistent with prior observations^40,69^, although the effect of frequency on the proportion of induced *habitual* seizures was smaller in our study (1.5-fold difference between high and low frequency) compared to prior reports in adults (e.g., 3-fold difference^40^).

### Yield: Suggestions for clinical practice

We interpret the larger magnitude of the frequency effect in our study to reflect a developmental difference in susceptibility to 1 Hz stimulation, such that 1 Hz stimulation seems to be particularly high-yield for inducing seizures in children. Consistent with that interpretation, we directly measured a significant interaction between age and frequency within our mixed-age cohort, highlighting that 1 Hz stimulation is particularly crucial to include when stimulating the *youngest* patients. Speculatively, these results further suggest that it may be fruitful to explore a broader range of stimulation frequencies for inducing seizures in the developing brain, a hypothesis we plan to test with future studies. We also saw a dramatic relationship between age and charge density, consistent with prior observations of decreased cortical excitability threshold with increased age during functional mapping^53,55^. This finding suggests that it is reasonable to expect to use higher current intensities in younger children and lower current intensities in older children and teenagers.

### Effects of anti-seizure medications and MRI lesion

ASM status and MRI lesion on the types of induced seizures. Although these effects were not the focus of the current study, an important observation is that seizures were induced regardless of ASM status, although the likelihood of an induced seizure being habitual electroclinical was higher for patients on ASMs as compared to off. We did not adjust this comparison for the patients’ baseline seizure frequencies, but typical practice in our center is that patients with higher baseline seizure frequencies are more likely to be maintained on their medications. The actionable conclusion is that clinicians ought not to worry too much about stopping (or restarting) a patient’s ASMs prior to ESIS; seizures can be obtained in either scenario, although habitual seizures may be more frequently obtained in patients on ASMs.

Regarding lesional status, prior work has shown that the presence of a lesion can increase the stimulation threshold; it has even been shown that lesion removal can cause the threshold to return to a lower level^70^. In contrast, we found that overall, patients with MRI lesions were just as likely as those without MRI lesions to have at least one induced seizure. There were, however, notable differences in the proportions of seizure types induced in each of those groups, with habitual electroclinical seizures accounting for the largest proportion of induced seizures in patients with lesions and the smallest proportion of induced seizures in patients without lesions. We are currently evaluating whether these between-group differences in induced seizure types depend more precisely on where the stimulation is applied, for example in an intralesional, perilesional or extralesional region. The current results nonetheless support approaching patients with and without MRI lesions similarly for ESIS.

### Induced seizure types

Another important novel observation from this study is our overall very high rate (89%) of induction of seizures of *any type*. This likely at least partially reflects our observation of an unexpectedly large number of induced *subclinical* seizures, something that has not been previously reported (although see^48^). We plan to evaluate, in the near future, the possible prognostic and/or localizing value of induced subclinical seizures, motivated by their high prevalence in our study (30% of all seizures) in combination with studies showing that *spontaneous* subclinical seizures are overall more common in children than adults^71–74^ and relate to surgical outcomes^73–75^. Speculatively, induced subclinical seizures obtained outside of the primary clinical SOZ may represent, for example, a marker of latent or future ictogenic potential, and thus be of prognostic value. This hypothesis is consistent with one prior study showing that induced subclinical seizures were seen in 14% of patients with good surgical outcomes and in 55% of patients with a poor surgical outcome, a significant difference^48^. Relatedly, for “cortical dysplastic regions” it has been shown that in resections guided by intraoperative electrocorticography, the completeness of the resection of regions showing repetitive electrographic seizures was correlated with surgical outcome^76^.

### Limitations and Future Directions

It would be ideal to understand more thoroughly how anatomic, methodologic and patient-specific variables and their interactions may predict seizure induction in pediatric patients with DRE. Although such analyses are beyond the scope of the current report, multivariable models of candidate predictors are presently being tested. We hope that such multivariable modeling will help define a more efficient and personalized approach to ESIS for children, recognizing that a truly comprehensive paradigm (incorporating two frequencies of stimulation at every single gray matter channel) may not be feasible for all busy EMU clinicians.

### Conclusions

Electrical stimulation for induction of seizures (ESIS) is an adjunctive tool that has long been employed to assist with surgical planning in adults. This prospective study of n=70 patients establishes that ESIS is safe, well-tolerated and high-yield in children and young adults with DRE undergoing sEEG. These novel results underscore that seizure stimulation can be safely and feasibly integrated into the clinical workflow in pediatric EMUs. Future directions include clarifying the relationship between induced seizures and surgical outcomes in children with DRE, specifically to determine ways in which the most accurate interpretation of induced seizures may depend on patient age.

## Data Availability

All data produced in the present study will be made available upon reasonable request to the authors, pending the completion of ongoing additional analyses including outcomes data. These ongoing analyses are expected to be completed by 2027.

---

Supplemental Table 1: Patient and session characteristics, stratified by whether or not patient had at least one *habitual* induced seizure

Appendix A: Thematic summary of pre-stimulation comments from parents and patients

Appendix B: Thematic summary of post-stimulation comments from parents and patients

## Acknowledgments

We are grateful to all the participants and their caregivers, without whose gracious involvement we would not have been able to conduct this study. We also thank all of our colleagues in the CCHMC Epilepsy Monitoring Unit who provided clinical care to the study participants during their sEEG admissions. We thank Madeleine Robben for assistance with data entry and management.

## Funding

This research was supported by the National Institute of Neurological Disorders and Stroke of the National Institutes of Health under award number K12NS098482.

ClinicalTrials.gov ID is NCT05469373.

## Conflicts of interest disclosures

None of the authors has any conflict of interest to disclose. We confirm that we have read the journal’s position on issues involved in ethical publication and that we affirm that our report is consistent with these guidelines.

## Notes

### Competing Interest Statement

The authors have declared no competing interest.

### Clinical Trial

NCT05469373

### Author Declarations

The Cincinnati Childrens Hospital Medical Center (CCHMC) Institutional Review Board (IRB) gave ethical approval for this work.

