## Supplemental Materials for "Electrical Stimulation for the Induction of Seizures is Safe, Well-tolerated and High-Yield in Children with Drug-Resistant Epilepsy"

### Supplemental Material

**Supplemental Table 1** Patient and session characteristics, stratified by whether the patient had at least one ***habitual*** **seizure** induced; for continuous variables, mean (SD); for categorical variables, number (proportion)

| **Patient and Session Characteristics, Continuous** | | | | | | |
|---|---|---|---|---|---|---|
| | **Overall Cohort (n=70)** | **Habitual Seizure Induced (n=46)** | **No habitual Seizure Induced (n=24)** | ***p* value** | **Hedge's g (effect size)** | ***t* statistic** |
| **Age at Stimulation, years** | 11.1 (SD 5.7) | 10.8 (SD 5.7) | 11.1 (SD 6.2) | 0.84 | 0.0489 | 0.1963 |
| **Age of Epilepsy Onset, years** | 4.4 (SD 4.4) | 4.2 (SD 4.2) | 4.8 (SD 4.8) | 0.62 | 0.1248 | 0.5013 |
| **Duration of Epilepsy, years** | 6.6 (4.2) | 6.7 (SD 3.8) | 6.4 (SD 4.9) | 0.74 | -0.0813 | -0.3264 |
| **Duration of sEEG, nights** | 6.2 (SD 5.0) | 5.7 (SD 3.6) | 7.0 (SD 7.0) | 0.32 | 0.2477 | 0.9948 |
| **Number of electrodes** | 14.3 (SD 3.9) | 14.1 (SD 3.7) | 14.6 (SD 4.4) | 0.61 | 0.1286 | 0.5164 |
| **Number of electrode contacts** | 158 (SD 48) | 157 (SD 45) | 161 (SD 56) | 0.77 | 0.0726 | 0.2915 |
| **Days between sEEG implant and stim.** | 2.9 (SD 2.9) | 3.2 (SD 3.3) | 2.4 (SD 2.0) | 0.29 | -0.2640 | -1.0601 |

| **Patient and Session Characteristics, Categorical** | | | | | | | |
|---|---|---|---|---|---|---|---|
| | **Overall Cohort (n=70)** | **Habitual Seizure Induced (n=46)** | **No habitual Seizure Induced (n=24)** | ***p* value (two-sided)** | **Test statistic*** | **Diff. (Δ) in proport-ions** | **Wald 95% CI for Δ, with Yates correction** |
| **Sedated on dexmedetomidine in ICU** | | | | | | | |
| Yes | 4 | 2 (0.50) | 2 (0.50) | 0.60 | 0.50 (FET) | 0.17 | (-0.80, 0.47) |
| No | 66 | 44 (0.67) | 22 (0.33) | | | | |
| **Spontaneous seizure <24 h prior to stim.** | | | | | | | |
| Yes | 47 | 31 (0.66) | 16 (0.34) | 1.00 | 1.03 (FET) | 0.01 | (-0.26, 0.28) |
| No | 23 | 15 (0.65) | 8 (0.35) | | | | |
| **Spontaneous seizure <24 h after stim.** | | | | | | | |
| Yes | 44 | 30 (0.68) | 14 (0.32) | 0.61 | 1.34 (FET) | 0.06 | (-0.20, 0.33) |
| No | 26 | 16 (0.62) | 10 (0.38) | | | | |
| **All spontaneous seizures AFTER stim.** | | | | | | | |

| | | | | | | | |
|---|---|---|---|---|---|---|---|
| Yes | 8 | 6 (0.75) | 2 (0.25) | 0.71 | 1.65 (FET) | 0.10 | (-0.29, 0.50) |
| No | 62 | 40 (0.65) | 22 (0.35) | | | | |
| **Laterality of sEEG implant*** | | | | | | | |
| Right | 20 | 15 (0.75) | 5 (0.25) | 0.53 | 1.76 (FET) | 0.12 | (-0.19, 0.43) |
| Left | 27 | 17 (0.63) | 10 (0.37) | | | | |
| Bilateral | 23 | 14 (0.65) | 9 (0.35) | | | | |
| **Medication Status**** | | | | | | | |
| On (full dose) | 17 | 13 (0.76) | 4 (0.24) | 0.79 | 1.30 (FET) | 0.06 | (-0.20, 0.32) |
| On (reduced dose) | 9 | 5 (0.56) | 4 (0.44) | | | | |
| Off | 41 | 26 (0.63) | 15 (0.37) | | | | |
| Mixed | 3 | 2 (0.67) | 1 (0.33) | | | | |
| **Sex** | | | | | | | |
| Female | 31 | 20 (0.65) | 11 (0.35) | 1.0 | 0.91 (FET) | 0.02 | (-0.27, 0.23) |
| Male | 39 | 26 (0.67) | 13 (0.33) | | | | |
| **Handedness***** | | | | | | | |
| Right-handed | 46 | 30 (0.65) | 16 (0.35) | 1.0 | 0.94 (FET) | 0.01 | (-0.31, 0.28) |
| Left-handed | 18 | 12 (0.67) | 6 (0.33) | | | | |
| Ambidextrous/Unknown | 6 | 4 (0.67) | 2 (0.33) | | | | |
| **Etiology** | | | | | | | |
| Lesional | 56 | 39 (0.70) | 17 (0.30) | 0.21 | 2.29 (FET) | 0.20 | (-0.14, 0.53) |
| Non-lesional | 14 | 7 (0.50) | 7 (0.50) | | | | |
| **Prior Epilepsy Surgery** | | | | | | | |
| Yes | 14 | 7 (0.50) | 7 (0.50) | 0.21 | 0.44 (FET) | 0.20 | (-0.53, 0.14) |
| No | 56 | 39 (0.70) | 17 (0.30) | | | | |

Counts represent 70 patient sEEG admissions, including three patients who had two distinct sEEG admissions.

FET = Fisher exact test; $\chi^2$ = Chi-squared test.

*For sEEG laterality groups, the right and left sided implants were compared using Fisher Exact Test, and the bilateral group was ignored.

** For ASM status, On (full dose) and On (reduced dose) were combined and compared against Off. The n=3 patients with mixed medication status (on meds for some trials, off meds for others) were ignored.

*** For handedness, right and left handed patients were compared using Fisher Exact Test, the small number of patients (n=6) in the ambidextrous/unknown group were ignored.

**Appendix A**
**Thematic Summary of Pre-Stimulation Comments from Parents and Patients**

1. Concerns About Seizures, Atypical Events, or Medical Risks
    - *“Worried about what it might feel like.”* — Patient
    - *“Only worried about prolonged seizures and possible need for rescue meds.”* — Parent
    - *“Hoping to avoid big seizures we saw last time.”* — Parent
    - *“Concerned we will see seizures not of his typical type.”* — Parent
    - *“Seizures are never fun — stimulated or spontaneous!”* — Parent
    - *“Concern with impact on developing brain.”* — Parent
    - *“Want to make sure we aren’t introducing new unknown affecters which could impact her sEEG.”* — Parent

2. Emotional Discomfort
    - *“A little nervous.”* — Patient
    - *“Nervous — hard to see your child have seizures.”* — Parent
    - *“New to us, so somewhat apprehensive.”* — Parent
    - *“Unknowns are always a little uncomfortable, but risks have been explained well.”* — Parent

3. Trust in Providers and Comfort with Process
    - *“Super comfortable.”* — Patient
    - *“I am trusting the neurologist’s information and thankful for the possibility of more information.”* — Parent
    - *“I think this will provide us good information.”* — Parent
    - *“I am very comfortable with stimulation (to find better results for epilepsy connections).”* — Parent

4. Desire to Contribute to Research
    - *“No, I am happy to help out with studies.”* — Parent
    - *“If data will be helpful to [my child’s] treatment.”* — Parent
    - *“Hope the data can be used both for research and for her individual treatment.”* — Parent

**Appendix B**
**Thematic Summary of Post-Stimulation Comments from Parents and Patients**

1. Continued Concerns About Seizures or Side Effects
   - *“Still a little scary.”* — Patient
   - *“Hand moves in weird waves.”* — Patient
   - *“Some of the feelings made me really uncomfortable. They didn't kill me! It was a very different experience.”* — Patient
   - *“Minimal. Seizures subsided faster than I expected.”* — Parent
   - *“I wasn’t sure if the stimulation contributed to the cluster seizures she experienced.”* — Parent
   - *“Good info but hate provoking seizure activity.”* — Parent

2. Discomfort Witnessing the Procedure
   - *“It was wholly unpleasant to witness the experimentation on my child.”* — Parent
   - *“Seeing the direct and immediate response he had to the push of a button made me feel like he was a kind of puppet.”* — Parent
   - *“Main concern is child’s perception of possible trauma.”* — Parent

3. Trust in the Process and Reassurance After the Procedure
   - *“There is still risk, but I feel comfortable. I felt safe.”* — Patient
   - *“Progressive ramp up to simulate seizure was good to ease comfort; VERY INFORMATIVE.”* — Patient
   - *“It was better than I thought.”* — Patient
   - *“Seems pretty safe. Did exactly what the test was supposed to do.”* — Parent
   - *“Test was run really well in my opinion. I appreciated all of the communication.”* — Parent
   - *“The doctor made us feel very comfortable about the testing.”* — Parent

4. Gratitude and Altruistic Motivation
   - *“Please do this to help doctor and other patients.”* — Patient
   - *“I have to do it to help me.”* — Patient
   - *“I think it is an excellent study to give more data.”* — Parent
   - *“Got good info – happy we did it!”* — Parent
